# An objective neurophysiological study of subconcussion in female and male high school student athletes

**DOI:** 10.1101/2024.03.20.24304610

**Authors:** Ryan C.N. D’Arcy, David McCarthy, Derek Harrison, Zander Levenberg, Julian Wan, Aidan Hepburn, Eric D. Kirby, Tanja Yardley, Nikita Yamada-Bagg, Shaun D. Fickling, Thayne A. Munce, David W. Dodick, Christopher Ahmad, Ken Shubin Stein

## Abstract

Emerging evidence from neurophysiological brain vital sign studies show repeatable sensitivity to cumulative subconcussive impairments over a season of contact sports. The current study addressed the need for a large prospective study comparing a low-contact control group to high-contact group. Importantly, the study also expanded the scope of neurophysiological changes related to repetitive head impacts to include female athletes in addition to male athletes. In total, 89 high school student athletes underwent 231 brain vital sign scans over a full calendar year. The results replicated prior subconcussive cognitive impairments (N400 delays) and sensory impairments (N100 amplitude reductions) in male athletes and demonstrated similar subconcussive impairments for the first time in female athletes. While there was no significant subconcussive difference between female and male athletes, female athletes show overall larger responses in general. The findings demonstrated that subconcussive impairments are detectable in a controlled experimental comparison for both female and male high school athletes. The study highlights the opportunity to monitor subconcussive changes in cognitive processing for both female and male athletes to help advance prevention, mitigation and management efforts aimed at reducing athletes’ risk of potential long-term negative health outcomes related to cumulative exposure to repetitive head impacts.

## Introduction

### Background

Head impact exposure from participating in contact or collision sports is commonly associated with concussion; however, subconcussion is a growing concern for athletes as well as a developing research area in parallel (Smith et al. 2019). Subconcussion has been increasingly implicated with potential short-term consequences on the developing brains and longer-term concerns related to chronic traumatic encephalopathy (McKee et al. 2009; Guskiewicz et al. 2005; Dioso et al. 2022). Concussion research has also historically been primarily focused on males (Valera et al. 2021; Snedaker et al. 2022). Therefore, emerging subconcussive research must be inclusive of both female and male athletes. This is a timely priority as new intervention options begin to emerge from the clinical trials literature (Breuer et al. 2023).

Operationally defined, a subconcussive impact is a mechanical force transmitted to the brain below the threshold for a diagnosis of an acute concussive injury. The effects of these repetitive head impacts (RHI) may neither be detectable to players nor to observers; however, it has been shown that RHI over time can result in both microstructural alterations and functional brain impairment (Mainwaring et al. 2018). The extent of impairment is highly correlated with the frequency of exposure to these impacts (Hirad et al. 2019; Saghafi et al. 2018; Fickling et al. 2019; Fickling, Smith, et al. 2021; Fickling, Poel, et al. 2021).

Recent research has increasingly focused on the role of cumulative RHI exposure rather than a concussion as a singular, acute traumatic event. Evidence suggests that even a single practice session involving head contact, such as heading a ball in soccer, can result in impairment (Nowak et al. 2020). Magnetic resonance imaging (MRI) studies have demonstrated that players exposed to RHI over the course of a season of sport show significant structural abnormalities compared to controls over the same time period (Hirad et al. 2019; Saghafi et al. 2018; Veksler et al. 2020). Both Hirad et al. (2019) and Saghafi et al. (2018) used diffusion-tensor imaging, an MRI-derived technique, to show abnormalities in the white matter of a group of football athletes were related to different measures of head impact exposure. Interestingly, while detecting differences in permeability and white matter microstructure in the brains of contact sport athletes, Veksler et al. (2020) did not find any statistical differences in blood-brain barrier permeability between non-contact athletes and non-athlete healthy controls, leading the authors to merge the groups into a single control group.

A major challenge for current medical imaging approaches, including MRI, relates to the need for accessible, objective, and sensitive measures of subconcussive impacts at points-of-care (PoC). Portable electroencephalography (EEG) provides the evaluation of individual neurophysiological event-related potentials (ERPs). These have recently been translated into a brain vital signs framework, facilitating access to sensitive measures for various PoC applications (Ghosh Hajra et al. 2016). Three well-established ERP responses are extracted within the brain vital signs framework: 1) the N100 for auditory sensation (Davis 1939); 2) the P300 for basic attention (Sutton et al. 1967); and 3) the N400 for cognitive processing (Kutas and Federmeier 2011). Brain vital signs have been successfully implemented in the evaluation of both concussion and subconcussion (Fickling et al. 2019; Fickling, Smith, et al. 2021; Fickling, Poel, et al. 2021).

Identification of subconcussive cognitive impairments over a season of contact sports was initially reported in male Junior-A ice hockey players [(Fickling et al. 2019), N=23]. Subsequent studies [(Fickling, Smith, et al. 2021), N=23; (Fickling, Poel, et al. 2021), N=15; (Breuer et al. 2023), N=30] replicated and confirmed identification of accumulated subconcussive exposure in male athletes for different age ranges (12-14, 16-21, & 21-28, respectively) and different contact sports, including ice hockey, tackle football, and mixed martial arts. Importantly, this work demonstrated strong, significant, and linearly predictive relationships between brain vital sign changes and exposure to subconcussive impacts (Fickling, Smith, et al. 2021; Fickling, Poel, et al. 2021). These relationships were demonstrated both when subconcussive impacts were quantified directly by impact sensors or indirectly by the number of games and practices played. A pattern of subconcussive changes have emerged, identified by changes in the N400 (cognitive processing) and N100 (sensory processing). The N400 and N100 components have been implicated in the rate of information processing and synchronous pyramidal neural activation (Luck 2014): the two are interdependent factors reflecting an association with cognitive processing (Kutas and Federmeier 2011) and auditory sensation (Davis 1939). While the replications of N400 and N100 subconcussive changes in male athletes are noteworthy, a single prospective study with a large sample size including female athletes is critical.

### Objectives & Hypothesis

The study was conducted at Brentwood College School (a Grade 8-12 high school in Mill Bay, British Columbia, Canada) by student researchers from the Brentwood Research, Action, and Innovation in Neuroscience (BRAIN) team. Brain vital signs were monitored in 89 female and male high school students (15-17 years of age) participating in high-contact (e.g. rugby, ice hockey, and soccer) and low-contact sports (e.g. rowing, climbing, and tennis) over three academic terms across the school year (Figure 1). With respect to matching participants in high-contact versus low-contact sports, Brentwood is a private boarding school, in which all students follow a structured program with commonly matched ages, daily schedules, diets, academics, athletics, and sleep routines. The study used a mixed-model, repeated measures design evaluated across 3 terms before and after each of the three school year terms as well as after the summer break, resulting in 7 possible scan timepoints. The objectives were to replicate prior subconcussion findings in male athletes, as well as in female athletes, with the primary hypothesis predicting that brain vital sign subconcussive differences would be detectable in the comparisons for high-contact sports relative to low-contact sports. Subsequent analyses were conducted to explore specific differences between female and male subconcussive and overall neurophysiological results.

**Figure 1:**
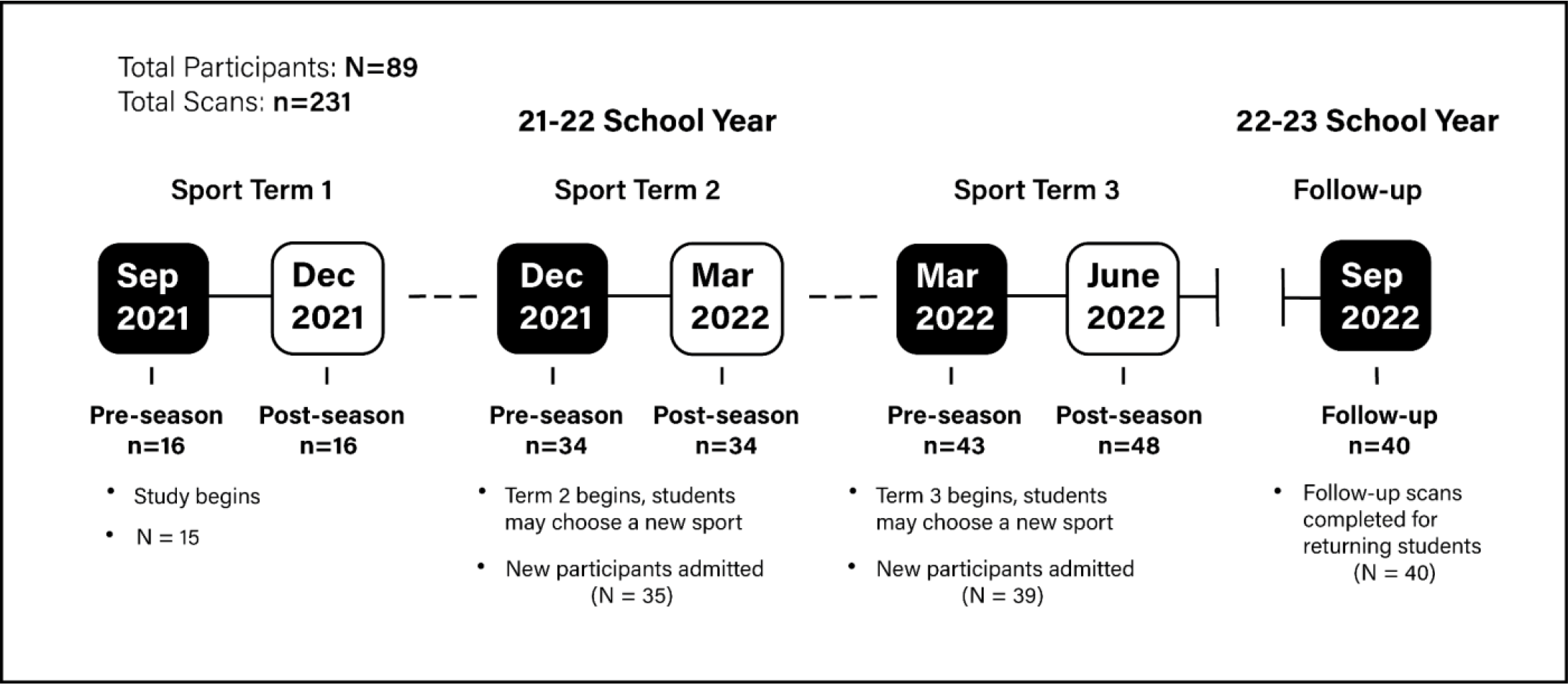
Overview of the experimental design and study timeline (N: Number of participants; n: brain vital signs scans completed).

## Results

In the current study, 89 female and male student athletes were scanned a total of 231 times before and after varying terms of contact or non-contact sports. All three brain vital signs, the N100, P300, and N400, were verified at both the group- and individual- levels. Peak amplitude and latency values were examined in the multivariate analysis of variance (MANOVA), which were first examined with univariate tests of normality and Levene’s test of equality of error variances. N100 latency and N400 amplitude failed tests of normality (*p* < 0.05), while all amplitude and latency values passed Levene’s test (*p* > 0.05). Therefore, non-parametric Kruskal-Wallis test results are also included in Supplemental Data 1 to demonstrate the robustness of the following MANOVA results.

### High-contact versus low-contact effects

Figure 2a shows brain vital sign differences in the N400 latency and N100 amplitude, while figure 2b provides the underlying waveform differences. The findings confirmed the consistent replication of subconcussive changes in the N400 and N100 at the overall group level, with a significant delay in N400 peak latency and a significant increase in N100 peak amplitude. Specifically, the MANOVA showed a significant main effect of condition between high- versus low-contact groups [*F*(6,104) = 7.95, *p* < 0.001, η_p_^2^ = 0.314] (Table 1). Separate univariate between-subjects tests on the outcome variables revealed significant effects of group on the N400 latency, [*F*(1) = 35.79, *p* < 0.001, η_p_^2^ = 0.247], and N100 amplitude, [*F*(1) = 7.88, *p* = 0.006, η_p_^2^ = 0.067] (Figure 2, Table 2). Figure 3 provides the individual scatter plot data for both the N400 latency and N100 amplitude effects, including means and standard deviations. The current results addressed the planned high- versus low-contact group comparison to build on prior pre- versus post- season differences in male athletes. Specific pre- versus post-comparisons are provided as Supplemental data for comparison (Supplemental Figure 1).

**Figure 2:**
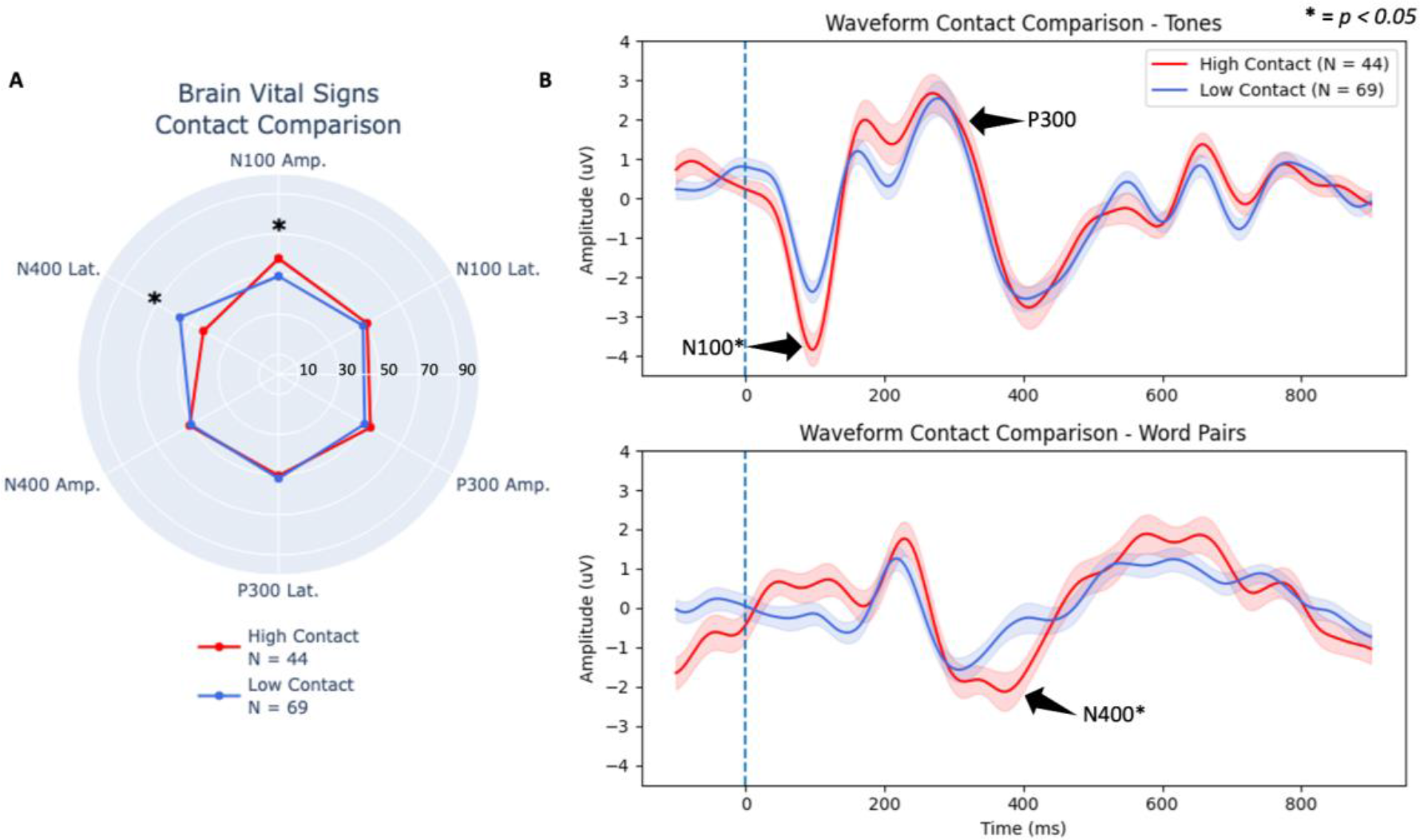
(A): Overall group (female and male combined) brain vital sign radar plot comparing high- versus low-contact groups. (B) Group-level grand average ERP waveforms from tones (top) and word pairs (bottom) for overall group comparing high- versus low-contact group differences. Deviant tones and incongruent word pair waveforms are plotted, with standard error shaded. N100, P300, and N400 brain vital sign components of the high-contact group are highlighted with labels and arrows. Asterisks denote significant effects in both the brain vital sign and waveform results.

**Figure 3:**
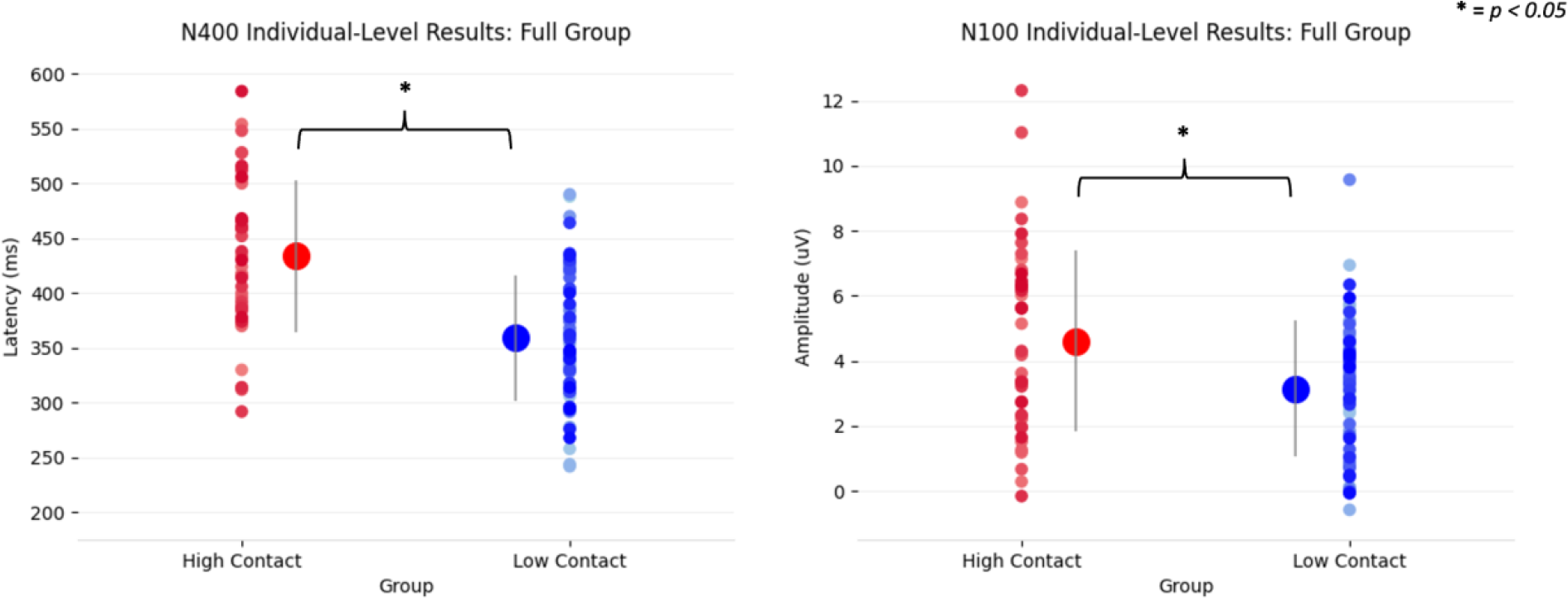
Individual scatter plot data, including group means and standard deviations (middle) showing a significant N400 latency and N100 amplitude differences for high-contact (red) versus low-contact (blue).

**Table 1:** MANOVA multivariate effect results for high- and low-contact scans.

| MANOVA | DF | PILLAI | F-VALUE | P-VALUE |
| --- | --- | --- | --- | --- |
| *GROUP* | 6,104 | 0.314 | **7.947** | **<0.001\*** |
| *SEX* | 6, 104 | 0.148 | **3.020** | **0.009\*** |
| *GROUP X SEX* | 6, 104 | 0.003 | 0.047 | 1.000 |

**Table 2:** MANOVA results for the group effect on each brain vital sign component.

| Brain vital sign | High-contact<br>mean (standard<br>deviation) | Low-contact<br>mean (standard<br>deviation) | *F*-value | *p*-value |
| --- | --- | --- | --- | --- |
| *N100 Amplitude* | -4.60 (2.82) | -3.15 (2.11) | **7.883** | **0.006\*** |
| *N100 Latency* | 100.78 (16.58) | 100.78 (25.64) | 0.750 | 0.388 |
| *P300 Amplitude* | 4.52 (2.84) | 3.86 (3.26) | 0.508 | 0.477 |
| *P300 Latency* | 274.77 (40.03) | 270.14 (35.22) | 0.304 | 0.583 |
| *N400 Amplitude* | -3.49 (2.85) | -3.35 (2.26) | 0.001 | 0.977 |
| *N400 Latency* | 433.50 (70.42) | 359.16 (57.86) | **35.786** | **<0.001\*** |

### Female compared to male subconcussive effects

There was no significant sex by contact group interaction present [*F*(6,104) = 0.047, *p* = 1.00, η_p_^2^ = 0.003] (Table 1), indicating female and male athletes experienced similar subconcussive effects. Figure 4a shows brain vital sign radar plots for female and male athletes for high- versus low-contact and figure 4b provides the corresponding waveform results. The MANOVA confirmed a significant main effect for female athletes compared to male athletes [*F*(6, 104) = 3.02, *p* = 0.009, η_p_^2^ = 0.148] (Table 1) across these scans. Separate univariate between-subjects tests revealed significant effects for the P300 amplitude [*F*(1) = 7.45, *p* = 0.007, η_p_^2^ = 0.064] and N400 amplitude [*F*(1) = 4.71, *p* = 0.032, η_p_^2^ = 0.041] (Table 3). Note in Figure 4b that the vertical y-axis (Amplitude) for female athletes and male athletes is fixed, highlighting the overall increased response amplitude in female responses relative to male responses across all peaks.

**Figure 4:**
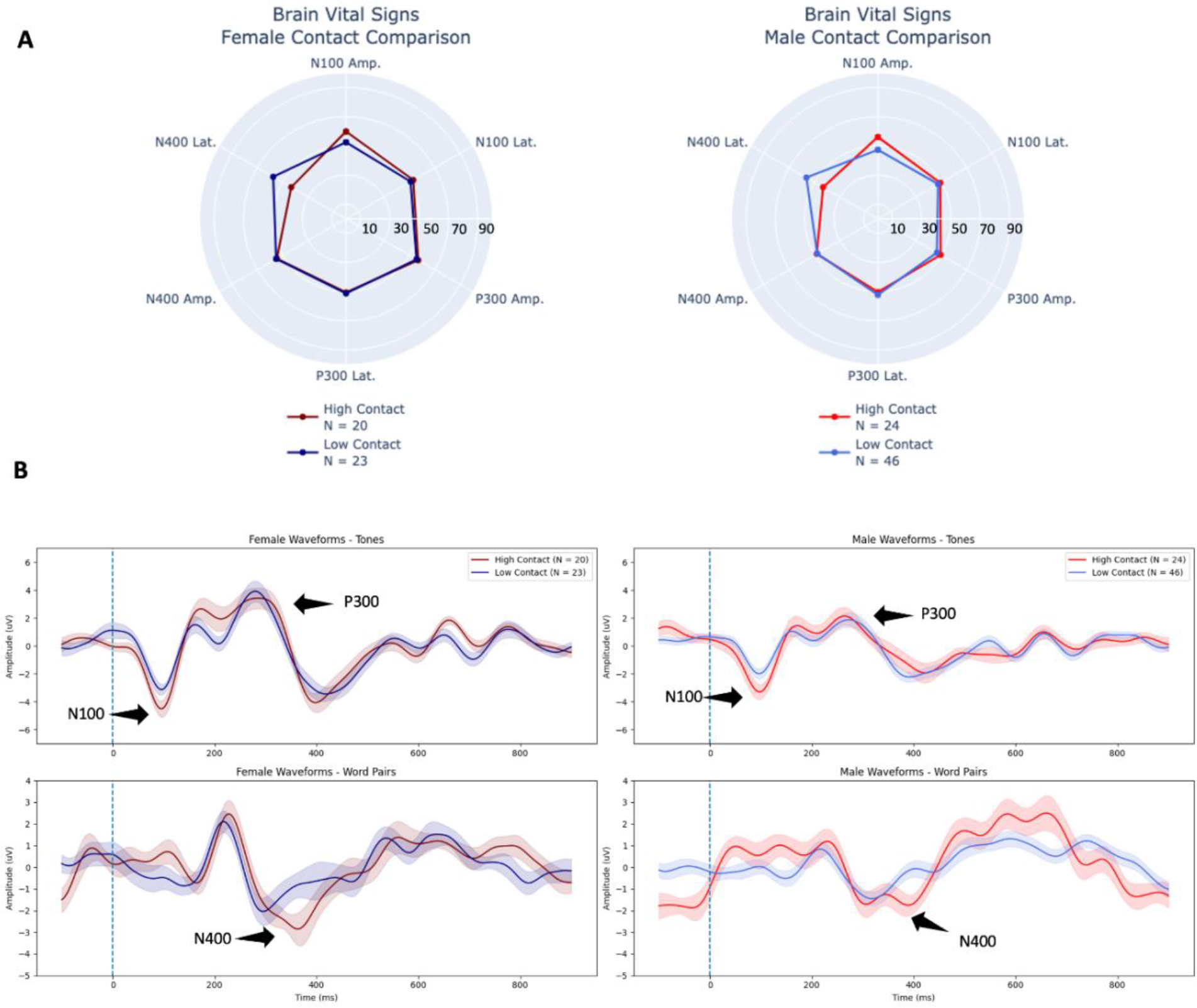
(A) Radar plots comparing individual-level brain vital signs of the high-contact and low-contact groups for both female and male athletes. **(B)** Group-level grand average ERP waveforms from tones (top) and word pairs (bottom) for female athletes (left, dark shading) and male athletes (right, light shading) comparing high- versus low-contact group differences. Deviant tones and incongruent word pair waveforms are plotted (with standard error shading). N100, P300, and N400 brain vital sign components of the high-contact group are highlighted with labels and arrows. Note in Figure 4b that the vertical y-axis (Amplitude) for female athletes and male athletes is fixed, highlighting the overall increased response amplitude in female responses relative to male responses across all peaks.

**Table 3:** MANOVA results for the main effect of sex on each brain vital sign component (high- and low-contact scans).

| Brain vital sign component | Female athlete mean (standard deviation) | Male athlete mean (standard deviation) | F-value | p-value |
| --- | --- | --- | --- | --- |
| N100 Amplitude | -4.29 (2.47) | -3.36 (2.47) | 2.427 | 0.122 |
| N100 Latency | 94.28 (15.32) | 101.86 (25.74) | 2.597 | 0.110 |
| P300 Amplitude | 5.18 (3.24) | 3.47 (2.85) | **7.447** | **0.007\*** |
| P300 Latency | 273.21 (37.50) | 271.17 (37.04) | 0.025 | 0.874 |
| N400 Amplitude | -4.07 (2.96) | -3.00 (2.08) | **4.707** | **0.032\*** |
| N400 Latency | 391.67 (78.34) | 385.91 (69.232) | 0.053 | 0.819 |

### Female compared to male effects

To further characterize brain vital sign profiles, the multivariate main effect of sex, and the univariate results above (Table 3) for female and male athletes, contact condition was collapsed across high- and low-contact scans to create Figure 5. P300 and N400 amplitude differences accounts for a significant main effect with a near significant effect in the N100 amplitude. The trends driving the main effect can be seen in Figure 6, which showed the individual scatter plots across female athletes and male athletes, including means and standard deviations. To further characterize female and male effects with a larger and closely weighted group number, Figure 7 provides radar plot and waveform results from female and male athletes for all collected scans (N=202 scans). The purpose of this comparison was to include all scans as an evaluation of robust differences between female and male athletes. Corresponding statistics are in Supplemental Data 3.

**Figure 5:**
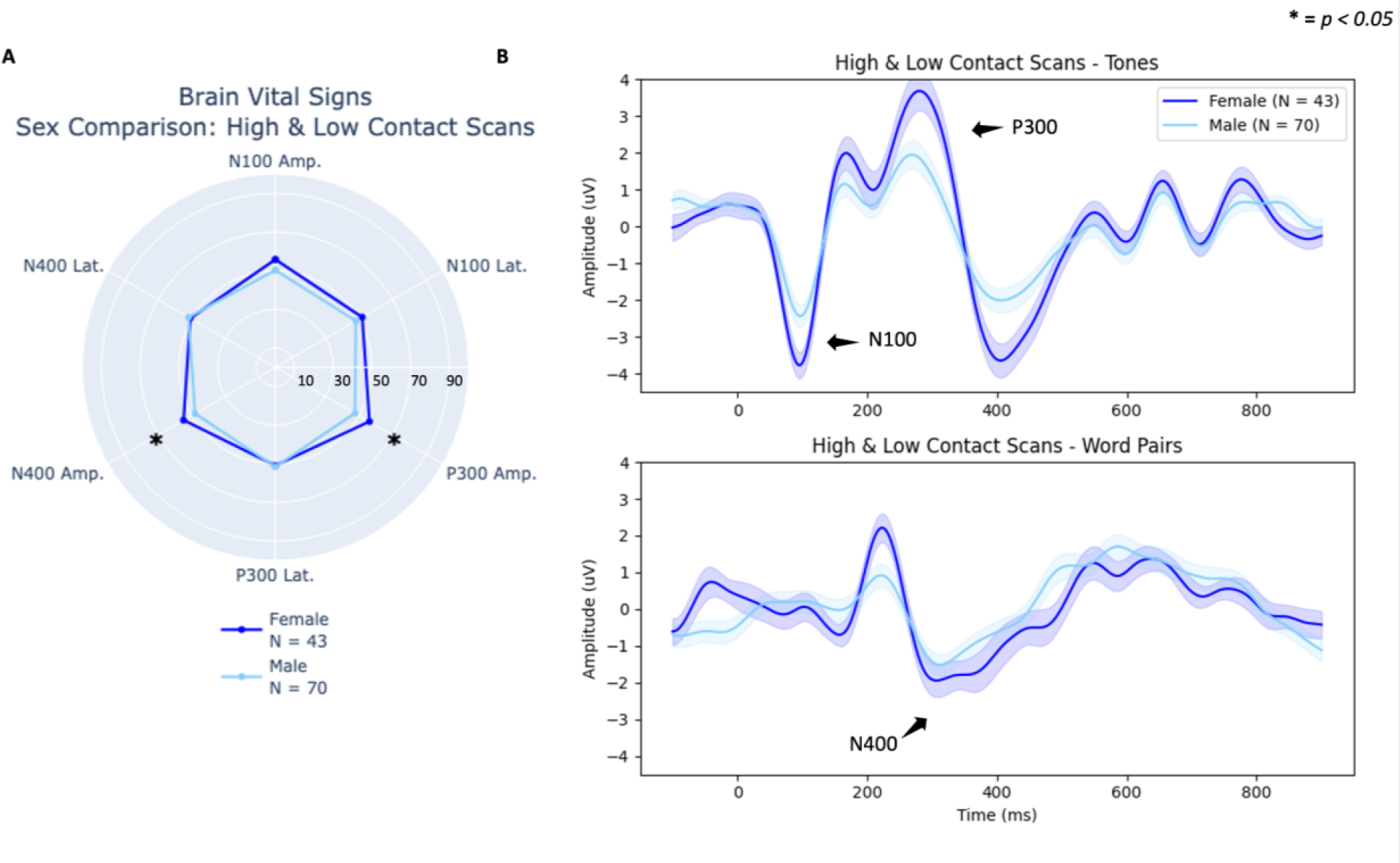
Comparison of all female and male high- and low-contact scans showing amplitude differences. **(A)** Radar plot showing overall female and male brain vital sign difference. **(B)** Group-level waveforms from deviant tones (top) and incongruent word pairs (bottom) are plotted (standard error shading) for female athletes (dark blue) and male athletes (light blue). N100, P300, and N400 components are highlighted with arrows.

**Figure 6:**
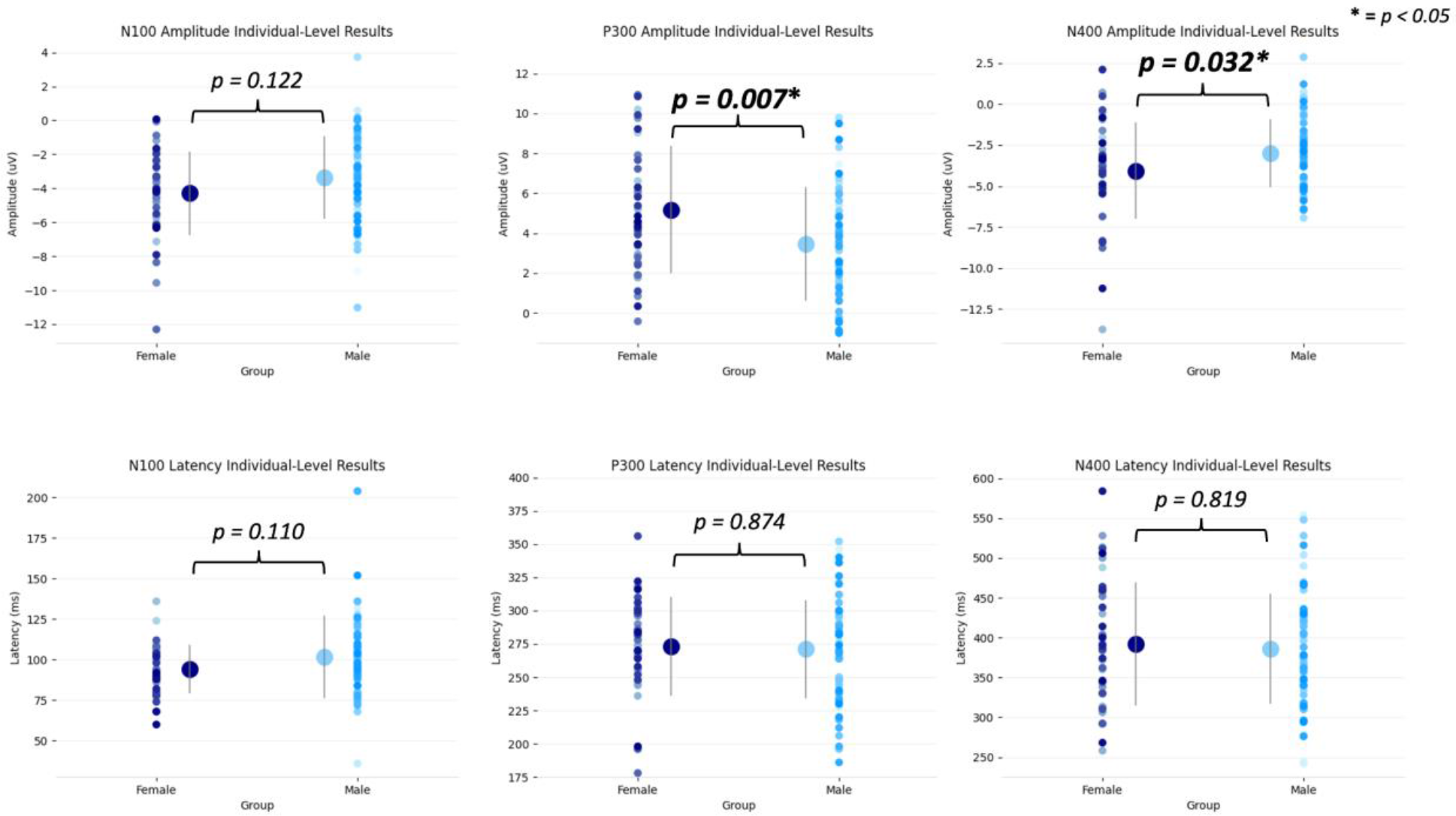
Scatter plots of individual-level data points for all scans of the amplitudes (top) and latencies (bottom) for all brain vital signs components in female athletes (left) and male athletes (right). Significant differences are highlighted by the asterisk, all *p*-values are reported as well (Bonferroni adjusted).

**Figure 7:**
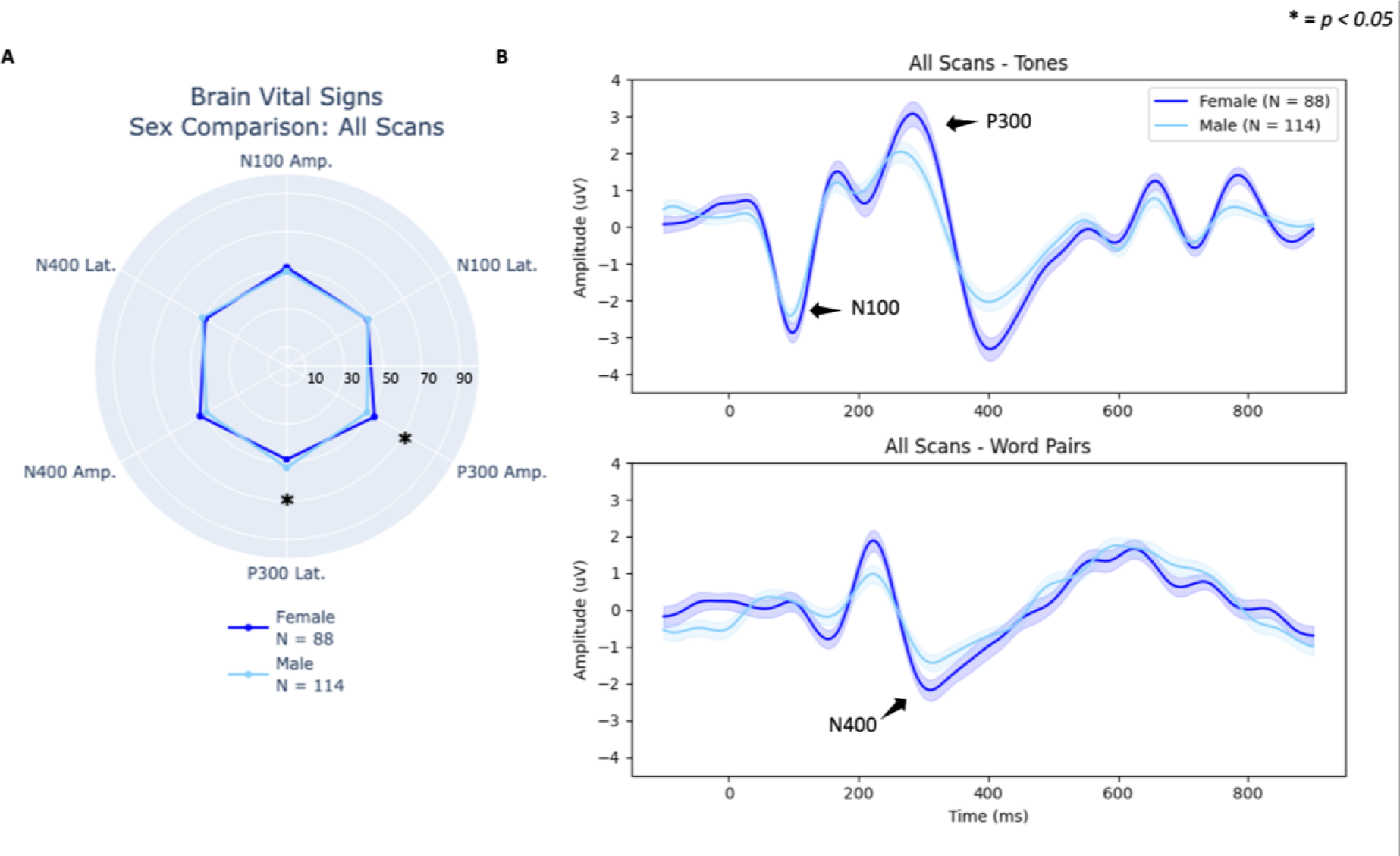
Comparison of all female and male scans. **(A)** Radar plot showing individual-level brain vital sign differences across all six components. **(B)** Group-level waveforms from deviant tones (top) and incongruent word pairs (bottom) are plotted (standard error shading) confirming lower amplitudes in male athletes (highlighted with arrows).

The current analyses examined high- versus low-contact, using equivalent exposure durations to prior studies. For additional analyses (N=202 scans), contact exposure levels were compared between low-contact exposure and both a single term and two or more terms of high-contact exposure. These results were closely consistent with both subconcussive and sex effects above and are provided as Supplemental Data 4.

## Discussion

The current findings replicated prior studies of subconcussive changes in brain vital signs. The study used a large, prospective, repeated measures design to compare high- versus low-contact exposure across both female and male athletes. The findings supported the primary hypothesis that prior subconcussive differences in the N400 and N100 would be detectable in a high- versus low- contact comparison (Figure 2), not only for male athletes but also in female athletes (Figure 4). Additional comparisons of female and male results showed significantly larger P300 and N400 responses for female athletes (Figure 6).

### Subconcussive analyses

Subconcussive N400 changes replicate four previous studies on subconcussion (Breuer et al. 2023; Fickling et al. 2019; Fickling, Smith, et al. 2021; Fickling, Poel, et al. 2021). The N400 is commonly associated with high-order cognitive semantic processing (Kutas and Federmeier 2011; D’Arcy et al. 2004) and measures changes in semantic processing, suggesting the initial demonstration of cognitive changes due to contact (Connolly and D’Arcy 2000; D’Arcy et al. 2003; Gawryluk et al. 2010; Ghosh Hajra et al. 2018). It is noteworthy that these replications have spanned across different sports and age ranges in male athletes. However, the specific N400 changes have varied with respect to latency delays and/or amplitude reductions, with the former directly measuring the rate of information processing and the latter related to synchronous pyramidal neural activation (Luck 2014). The two are interdependent factors reflecting a relative impairment in cognitive processing. While the initial results for female athletes showed comparable N400 latency delays, a specific pattern of change in N400 latency versus amplitude remains to be characterized.

Similarly, subconcussive changes in the N100 have been replicated across a number of prior studies. While the studies have reported only N100 amplitude changes, the direction of the effects has varied (i.e., decreases, increases, and correlated variability (Fickling et al. 2019; Fickling, Smith, et al. 2021; Fickling, Poel, et al. 2021). In prior linear regression analyses of the relationship between brain vital signs and helmet mounted accelerometers, inclusion of the N100 with the N400 provided additional explanatory variance (Fickling, Poel, et al. 2021). Consequently, N100 changes suggest that subconcussive impairments can span from lower-level sensory processing to higher level cognitive processing. As with the N400 above, the common preliminary results showing N100 changes in both female and male athletes do not rule out the possibility of specific differences. Indeed, examination of the female and male waveforms shows early sensory and attention differences in other ERP components (see below).

Of note, the current findings are the first to compare brain vital signs between high- and low-contact groups using a between-subjects design. To anchor to prior studies, a direct evaluation of the within-subjects pre- versus post- design has also demonstrated a significant delay in the N400 latency (Supplemental Data 2). The contribution of a control group is an important step, with the interpretation further underpinned by the nature of the closely matched comparison (i.e., related to questions about whether the effects are indeed attributable to subconcussive impact exposure). The student athletes in this study attended a private school with full time residence. Accordingly, all students follow a highly structured daily and weekly schedule, matched across programs, activities, diet, sleep, physical fitness, rest, and related factors.

Another novel aspect of the current study was the ability to begin exploring differences between a single term versus two or more terms of exposure to subconcussive impacts (Supplemental Data 4). While preliminary, the findings suggested that there may be a graded effect of exposure on the N400 peak latency (Supplemental Data 4). These results highlighted important questions related to tracking the extent of exposure with increased temporal resolution to characterize the changes over time. As reflected in the study design (Figure 1), a large cohort of these students were also re-evaluated after the summer break to determine whether this duration of rest resulted in a return to pre- baseline levels following post- subconcussive changes. Evaluation of these results are currently being analyzed as a follow-up study. While the emerging subconcussive findings are concerning, the demonstrated utility of using brain vital signs to objectively identify acute cognitive changes associated with repeated impact exposure has enabled clinical trials on potential interventions. A recent study on subconcussion focused on a prospective randomized placebo-controlled trial evaluating the effect of a dietary supplement effects created to reduce neuroinflammation and support energy production, on brain structure and function in Junior-A ice hockey players (Breuer et al. 2023). Breuer et al. (2023) randomized players into two groups: a group that took a supplement containing a combination of non-pharmacological ingredients shown to support brain health (Synaquell™ - Thorne Research Inc., NY, USA) daily over the season and a placebo control group who followed the same protocol without supplementation. Multivariate analyses showed significant neurocognitive pre- to post- season changes in only the control group, with significant negative impacts on the N100 and N400 along with reduced saccadic eye movement (King-Devick Test) and neurofilament light chain measures.

### Female and male comparison

Robust P300 and N400 differences between female and male athletes were shown with and without subconcussive contact exposure as a factor. While the significant P300 and N400 differences demonstrated larger amplitudes for female athletes, examination of the overall waveforms showed this finding generally across all ERP responses. There are at least two potential factors related to interpretation of the difference between female and male athletes. One factor may relate to general attention difference during the approximately 6-minute test time, whereby female athletes showed increased sustained attention on average relative to male athletes. In this case, improved sustained attention to the task would result in a higher signal-to-noise ratio in the signal averaged ERPs and consequently higher response amplitudes. Alternatively, another factor may relate to a fundamental difference in ERP response size between female and male individuals. While these two factors are not mutually exclusive, the notable differences in baseline variance suggested sustained attention during the test time is likely a factor (Figure 5). Differences in attention between female and male participants has been noted in prior literature, but these results are still mixed (Davidson, Cave, and Sellner 2000; Solianik, Brazaitis, and Skurvydas 2016; Riley et al. 2016).

As mentioned above, despite the subconcussive similarities between female and male athletes, it is too early to exclude differences, as an “absence of evidence is not evidence of absence.” For instance, examination of the ERP waveforms (Figure 4) shows noteworthy subconcussive differences in responses outside of the N100, P300, and N400. In particular, there appears to be a subconcussive reduction in the N200-P200 difference in female athletes, which is not present in male athletes. The N200-P200 is associated with early perceptual-attentional processing (Portella et al. 2012; D’Arcy, Connolly, and Crocker 2000; S. H. Patel and Azzam 2005), and further research should explore this in the context of sex-specific changes in subconcussion and head-impact exposure. With respect to this, the clinical importance of even small attentional impairments can consequently have a larger impact on high-level processing for individuals in attention demanding professions. For example, we identified early on in clinical concussion evaluation that these changes can have a compounding effect on high demand jobs such as air traffic controllers relative to less attentionally-demanding positions (Mateer and D’Arcy 2000). A comparable impact may be a consideration for enhanced sustained attention processing in female athletes. Given recent advances in understanding functional neuroanatomical differences (Andrushko et al. 2023), further studies needed to better characterize female- versus male- specific subconcussive profiles.

### Limitations and Future Directions

The current study included data across a large sample made up of smaller subsets across different sports, with differing contact levels. While this was beneficial to create a large sample size for this study, larger subsets of groups pertaining to each specific sport would help elucidate sport-specific differences. Similarly, history of high-contact sport participation and concussion are important variables that may affect the current state of participants. Nonetheless, the current study is the first in a continuing investigation that will subsequently focus on specific sports and recovery times following subconcussive impairment in contact sport. Future studies will expand to focus on recovery effects and differences following summer break. Also, the current study combined subjects in grades 10-12 in a highly structured school environment; however, previous work has shown differing neurological developmental rates between female and male adolescents (Paus 2010; Kaczkurkin, Raznahan, and Satterthwaite 2019; Gur and Gur 2016), which may have been a factor for female and male-specific profiles. Future work will include larger age subset sizes and longitudinal analyses, as well as controlling for age as a co-variate in future models, to have a better understanding of developmental differences between female and male individuals. Finally, the current study did not track important biological factors that differ between female and male athletes. These factors include but are not limited to menstrual cycle phase at time of exposure or in the case of multiple exposures, when the majority of exposures occurred (menstrual phase has been a predictor of mild TBI outcome (Wunderle et al. 2014) and progesterone shows neuroprotective as a TBI treatment (Wright et al. 2007)); body fat percentages given recent understanding of fat being a major endocrine organ (Musi and Guardado-Mendoza 2014); and diet given the understanding of diet impact on inflammation and brain function (P. R. Patel et al. 2023; Markovic et al. 2021). With clear neurophysiological differences identified between females and males, future work must include expanded characterization of these key biological factors to understand change that may affect results at the time of evaluation. Lastly, to better understand the resulting effect of these subconcussive neurophysiological changes and differences, additional study variables should include heart rate variability data to understand clinical and sub-clinical autonomic dysfunction and time to recovery, as well as changes in sleep and endocrine levels due to RHI, as TBI may lead to endocrine changes, specifically in developing adolescents (Richmond and Rogol 2014).

## Conclusion

With increasing attention on subconcussive impairments and potential interventions to limit future risks (e.g., long-term neuropsychiatric impairment, chronic traumatic encephalopathy – CTE), the current findings provide a large sample demonstration of subconcussive changes between closely matched high- versus low-contact student athletes that replicate previous investigations. The findings provide strong support for continued focus on the role of subconcussive injuries in sport-related brain health research. Importantly, as this field of study has historically focused on male athletes, inclusion of female athletes in the current findings represents a critical milestone towards understanding specific neurophysiological differences before and after exposure to subconcussive impacts. Initial results strongly support continued investigations that span beyond male athletes in helmeted contact sports.

## Methods

A total of 89 high school students were scanned 231 times over the year (34 females and 55 males). In total, 8 scans were removed due to attrition, 12 scans had a data collection error, failing to produce scan data, 5 due to dropping all epochs for a condition (i.e. deviant tones), and 4 due to excess noise (N=202 scans). For analysis and results we ended with N=81, n=202 (33 females and 48 males). The current study focused on a scan-based between contact groups analysis, instead of a pre-post subject based within-subjects analysis. I.e. results from a subject scan after a season of low contact sport would be characterized as a low contact data point (not a benchmark or pre data point), while if that same subject had a separate scan later in the school year after 2 terms of high contact, that scan would be characterized as a high contact data point. This was done to maximize the total data point number for analyses and provide more equal weighting across groups, as opposed to a repeated measure subject-based analysis including only subjects that had not had any contact immediately prior to the study.

All healthy participants were in grades 10 through 12 (age range: 15-17), fluent in English, no history of concussion in the past 6-months, no self-reported problems with brain function, and had no hearing problems. All participants were recruited from Brentwood College School athletic programs, which involve either high-contact (e.g., rugby, ice-hockey, soccer, field hockey, jujitsu, and basketball) or no-contact/low-contact sports (e.g., rowing, tennis, and climbing) over three distinct three-month terms in the school year (Fig. 1). As described above, Brentwood is a private boarding school in which all students follow a structured daily schedule and a similar diet, exercise, and sleep schedule throughout the academic year. Accordingly, participants were closely matched across a wide number of typical lifestyle factors. The study had Advarra institutional research ethics board approval and all participants along with their guardians provided informed consent.

### Data Collection

The brain vital sign framework (Ghosh Hajra et al. 2016) was employed with the NeuroCatch® Platform (Version 1.1), which uses EEG to rapidly extract the N100, P300, and N400 ERPs. All three ERPs were stimulated using a compressed and standardized auditory stimulus sequence (approximately 6 minutes) consisting of sets of tones and spoken word pairs. The brain vital signs framework utilizes an oddball paradigm, involving frequent standard tones with infrequent oddball tones, which elicit the N100 and P300 components. The tone stimuli precede word pairs, which are either matched or mismatched semantically-related word-pair primes (e.g., match: bread-butter; mismatch: bread-window) to elicit the N400 component. Participants passively listened to the stimuli and decided if the word pairs matched or mismatched.

Participants were fitted with an elasticized cloth cap containing an 8-channel EEG amplifier with standardized electrode locations (Fz, Cz, & Pz). Three additional electrodes were placed on the participant’s forehead to record the ground (GND) and electrooculogram (vEOG, hEOG) signals. A reference electrode was clipped to the participant’s right earlobe. Standard EEG skin preparation (70% isopropyl alcohol wipes) and conductive gel products (SignaGel) were used to ensure appropriate contact for all EEG sensors. Distractions were mitigated by performing the scans in a quiet and closed classroom.

The same environment was used for all scans. Participants were asked to listen attentively to the stimuli, but no active response was required. To reduce motor and ocular artifacts, participants were instructed to sit motionlessly, and maintain visual fixation on a cross positioned at eye level 2 m away.

### Data Processing

The NeuroCatch® Platform was calibrated for any trigger latency delays. The most common delay was identified and applied to shift all trigger data. EEG data were processed using a fourth order Butterworth filter (0.5-10 Hz) and a Notch filter (60 Hz). Adaptive filtering using electrooculogram channels was used to correct for ocular artifacts. EEG was epoched based on stimulus events using an epoch from -100ms pre-stimulus to 1000ms post-stimulus. All epochs were linearly detrended and any epochs with noise exceeding ±75μV were rejected. N100, P300, and N400 peaks were calculated using the maximal peak within a standard temporal window and manually verified. Amplitude and latency metrics from these peaks were then linearly transformed into standardized scores on a scale from 0 to 100, derived from entire group means (Fickling et al. 2019). All preprocessing was completed in Python using the Scipy and MNE libraries.

### Statistical Analysis

All individual-level statistical analyses of peak amplitude and latencies were performed using SPSS (Version 29.0.0, IBM, NY, USA). In the current study, individual-level can be defined as amplitudes and latencies from each individual scan data, not derived from the group-level average waveforms. To assess differences in brain vital signs between contact groups and sex, brain vital sign scores were compared using a multivariate analysis of variance (MANOVA) (**Between:** Group: low-contact and high-contact; Sex: Female and Male) on N100, P300, and N400 amplitudes and latencies for only scans characterized as low-contact or high-contact for two or more terms. To further assess differences in brain vital signs between sex in a larger number of scans, brain vital sign scores were compared using a multivariate analysis of variance (MANOVA) (**Between:** Sex: Female and Male) on N100, P300, and N400 amplitudes and latencies for all 202 scans collected. All univariate results are adjusted for multiple comparisons using Bonferroni adjustment. All statistics were also rerun only with subjects with less than 50% of epochs dropped (12 of 202 scans dropped) due to the 75μV thresholding measures mentioned above. To keep our participant number large and because the analysis resulted in very similar outcomes, the current manuscript reports the statistics from the full group data, including subjects with more than 50% of epochs dropped.

## Data Availability

All data are available from the corresponding author upon reasonable request, subject to adherence to the ethical guidelines and privacy policy governing this research.

## Acknowledgements

The authors would like to acknowledge the important leadership of Dr. Aynsley Smith (Mayo Clinic, retired), whose passionate leadership for research advances in sports-related concussion and ever-lasting priority to translate research advances to young females and males served as an important inspiration for this study. The authors would also like to acknowledge the technical assistance by Joshua Ighalo and the assistance in data collection by Laila Reed.

## Funding

This work has been supported in part by funding from the Natural Sciences and Engineering Research Council of Canada (Discovery Grant, PI R.C.N. D’Arcy; CGS M, E.D. Kirby).

## Competing interests

Some of the authors (EDK, TY, NYB, SDF, RCND) have financial and/or business interests in HealthTech Connex Inc., which may be affected by the research reported in this paper. Over the past 36 months, DWD reports the following competing interests: Consulting: Amgen, Atria, CapiThera Ltd., Cerecin, Ceruvia Lifesciences LLC, CoolTech, Ctrl M, Allergan, AbbVie, Biohaven, GlaxoSmithKline, Lundbeck, Eli Lilly, Novartis, Impel, Satsuma, Theranica, WL Gore, Genentech, Nocira, Perfood, Praxis, AYYA Biosciences, Revance, Pfizer. Honoraria: American Academy of Neurology, Headache Cooperative of the Pacific, Canadian Headache Society, MF Med Ed Research, Biopharm Communications, CEA Group Holding Company (Clinical Education Alliance LLC), Teva (speaking), Amgen (speaking), Eli Lilly (speaking), Lundbeck (speaking), Pfizer (speaking), Vector Psychometric Group, Clinical Care Solutions, CME Outfitters, Curry Rockefeller Group, DeepBench, Global Access Meetings, KLJ Associates, Academy for Continued Healthcare Learning, Majallin LLC, Medlogix Communications, Medica Communications LLC, MJH Lifesciences, Miller Medical Communications, WebMD Health/Medscape, Wolters Kluwer, Oxford University Press, Cambridge University Press. Non-profit board membership: American Brain Foundation, American Migraine Foundation, ONE Neurology, Precon Health Foundation, International Headache Society Global Patient Advocacy Coalition, Atria Health Collaborative, Arizona Brain Injury Alliance, Domestic Violence HOPE Foundation/Panfila. Research support: Department of Defense, National Institutes of Health, Henry Jackson Foundation, Sperling Foundation, American Migraine Foundation, Henry Jackson Foundation, Patient Centered Outcomes Research Institute (PCORI). Stock options/shareholder/patents/board of directors: Aural analytics (options), Axon Therapeutics, ExSano (options), Palion (options), Man and Science, Healint (options), Theranica (options), Second Opinion/Mobile Health (options), Epien (options/board), Nocira (options), Matterhorn (shares/board), Ontologics (shares/board), King-Devick Technologies (options/board), Precon Health (options/board), AYYA Biosciences (options), Axon Therapeutics (options/board), Cephalgia Group (options/board), Atria Health (options/employee). Patent 17189376.1-1466:vTitle: Onabotulinum Toxin Dosage Regimen for Chronic Migraine Prophylaxis (Non-royalty bearing). Patent application submitted: Synaquell® (Precon Health).

## Author Contributions

RCND assisted with project study design, manuscript drafting, and acted as lab PI. DM contributed with data collection and project study design. DH contributed with data collection, data processing, and manuscript editing. ZL contributed with data collection and manuscript editing. JW contributed with data collection and manuscript editing. AH contributed with data collection, statistical analysis, and manuscript editing. EDK contributed with data processing, statistical analysis, and manuscript drafting. TY, NYB, SDF, TAM, DD, CA, and KSS contributed with manuscript editing.

## Notes

### Author Declarations

Advarra institutional research ethics board gave ethical approval for this work.

### Summary of Updates

Citations/references have been updated. The title has been updated.

