## Supplemental Data for "An objective neurophysiological study of subconcussion in female and male high school student athletes"

### 1) Non-Parametric Results

**Supplementary Table 1:** Kruskal-Wallis results for the group effect on each brain vital sign component.

| **Brain vital sign component** | ***Kruskal-Wallis* statistic** | ***p*-value** |
|---|---|---|
| N100 Amplitude | **5.914** | **0.015*** |
| N100 Latency | 0.624 | 0.430 |
| P300 Amplitude | 1.980 | 0.159 |
| P300 Latency | 0.004 | 0.951 |
| N400 Amplitude | 0.190 | 0.663 |
| N400 Latency | **26.553** | **<0.001*** |

**Supplementary Table 2:** Kruskal-Wallis results for the sex effect on each brain vital sign component.

| **Brain vital sign component** | ***Kruskal-Wallis* statistic** | ***p*-value** |
|---|---|---|
| N100 Amplitude | 3.404 | 0.065 |
| N100 Latency | 1.886 | 0.170 |
| P300 Amplitude | **6.894** | **0.009*** |
| P300 Latency | 0.615 | 0.433 |
| N400 Amplitude | 3.831 | 0.050 |
| N400 Latency | 0.010 | 0.920 |

### 2) Pre/Post Fickling et al. Recreation

To create a more exact replication of previous subconcussive literature, only male subjects that had no or low-contact sport the term before a high-contact sport were included (N = 13). Peak amplitudes and latencies were used in a within subjects (Timepoint [2]: baseline and endpoint) repeated measures multivariate analysis of variance (RM-MANOVA) to assess subconcussive effects on subjects. Post-hoc univariate results were Bonferroni corrected (6 brain vital signs, therefore divided by 6). The main multivariate effect of timepoint was insignificant [$F(6,7) = 3.354$, $p = 0.069$, $\eta_p^2 = 0.742$]; however, previous work showing N400 latency differences is replicated and highlighted in the univariate results below (Supplementary Table 3). Both individual-level (individual peaks - Supplementary Table 3) and group-level (group averaged waveform - Supplementary Figure 1) show similar results that reflect previous studies.

**Supplementary Table 3:** Repeated measures MANOVA results for the recreation of previous subconcussive effects with repeated measures on each brain vital sign component.

| Brain vital sign component | Baseline mean (standard deviation) | Endpoint mean (standard deviation) | *F*-value | *p*-value |
|---|---|---|---|---|
| N100 Amplitude | -3.16 (2.35) | -2.76 (0.98) | 3.355 | 0.092 |
| N100 Latency | 143.38 (36.97) | 143.38 (31.29) | 0.119 | 0.736 |
| P300 Amplitude | 4.57 (2.97) | 4.21 (2.62) | 0.007 | 0.937 |
| P300 Latency | 307.08 (47.76) | 329.85 (44.00) | 0.064 | 0.804 |
| N400 Amplitude | -3.33 (1.93) | -2.70 (1.57) | 0.108 | 0.748 |
| N400 Latency | 355.85 (84.76) | 442.92 (50.74) | **4.974** | **0.046*** |

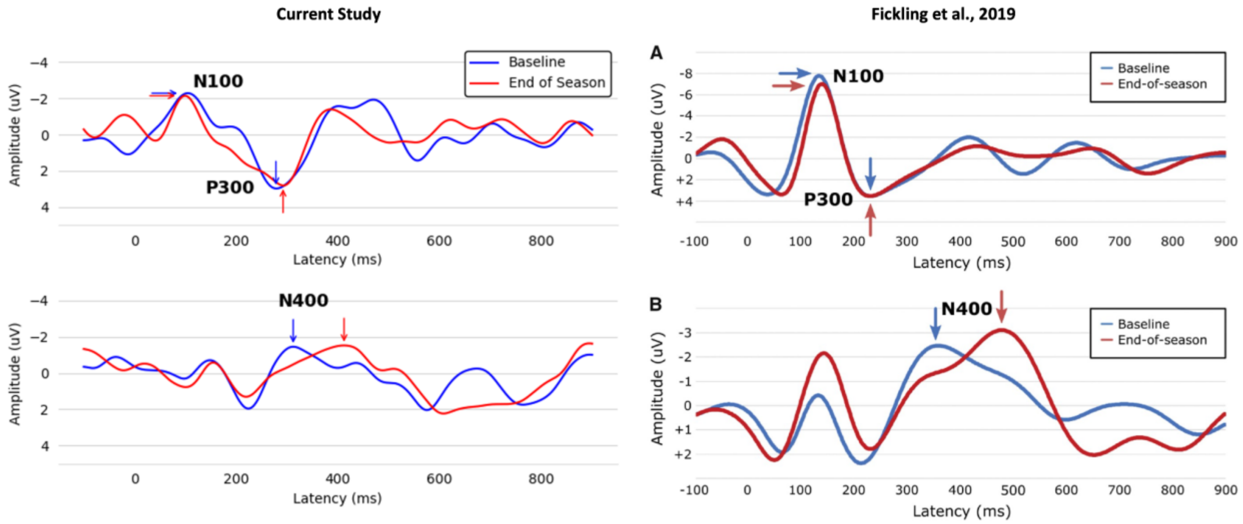


**Supplementary Figure 1:** Group-level waveforms from the current study repeated measures subset (left) and Fickling et al., 2019 (right) of deviant tones (top) and incongruent word pairs (bottom). Note negative amplitudes are up to make the comparison to the past Fickling et al., 2019 study easier. Right figure of Fickling et al., 2019 study was used with author's consent.

### 3) Full Group Sex Difference

The MANOVA confirmed a significant main effect for females compared to males [$F(6, 195) = 2.98$, $p = 0.008$, $\eta_p^2 = 0.084$]. Separate univariate between-subjects tests below (Supplementary Table 4).

**Supplementary Table 4:** MANOVA results for the main effect of sex on each brain vital sign component for all scans.

| **Brain vital sign component** | **Female mean (standard deviation)** | **Male mean (standard deviation)** | ***F*-value** | ***p*-value** |
|---|---|---|---|---|
| N100 Amplitude | -3.47 (2.58) | -3.21 (2.21) | 1.149 | 0.285 |
| N100 Latency | 101.28 (19.97) | 100.51 (24.99) | 0.040 | 0.842 |
| P300 Amplitude | 4.45 (2.93) | 3.58 (2.70) | **4.978** | **0.027*** |
| P300 Latency | 281.03 (37.39) | 267.60 (37.88) | **6.098** | **0.014*** |
| N400 Amplitude | -3.60 (2.63) | -3.05 (2.10) | 2.968 | 0.086 |
| N400 Latency | 401.46 (80.80) | 391.77 (69.78) | 0.731 | 0.393 |

### 4) Inclusion of single term high-contact exposure

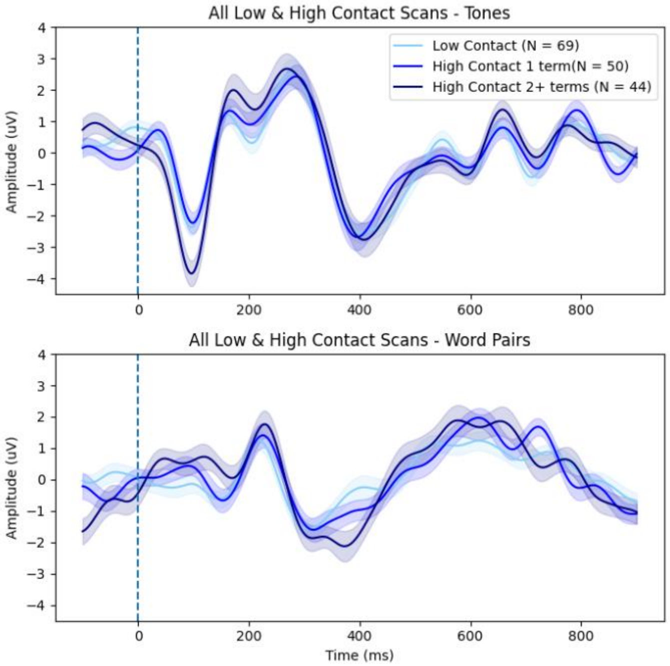


**Supplementary Figure 2:** Group-level waveforms from deviant tones (top) and incongruent word pairs (bottom) are plotted (standard error shading) for individuals who experienced low-contact, high-contact for 1 term, and high-contact for 2 or more terms to show graded effect of exposure.
